# *WDR12* and *HIVEP3* are Contributors to Cognitive Preservation in Amish SuperAgers

**DOI:** 10.64898/2026.02.02.26345224

**Authors:** Daniel Dorfsman, Michael B. Prough, Alex Gulyayev, Laura J. Caywood, Jason E. Clouse, Sharlene D. Herington, Susan H. Slifer, Larry D. Adams, Renee A. Laux, Yeunjoo E. Song, Audrey Lynn, Sarada L. Fuzzell, Sherri D. Hochstetler, Kristy Miskimen, Leighanne R. Main, Ping Wang, Yining Liu, Noel Moore, Paula Ogrocki, Alan J. Lerner, Jeffery M. Vance, Michael L. Cuccaro, Jonathan L. Haines, Margaret A. Pericak-Vance, William K. Scott

**Affiliations:** John P. Hussman Institute for Human Genomics, University of Miami Miller School of Medicine, 1501 NW 10^th^ Ave, Miami, FL 33136, USA; Dr. John T. Macdonald Foundation Department of Human Genetics, University of Miami Miller School of Medicine, 1501 NW 10^th^ Ave, Miami, FL 33136, USA; Department of Population and Quantitative Health Sciences, School of Medicine, Case Western Reserve University, 2013 Cornell Road, Cleveland, OH 44106, USA; Cleveland Institute for Computational Biology, School of Medicine, Case Western Reserve University, 2013 Cornell Road, Cleveland, OH 44106, USA; Department of Neurology, School of Medicine, Case Western Reserve University, 10900 Euclid Ave, Cleveland, OH 44106, USA; University Hospitals Cleveland Medical Center, 11100 Euclid Ave, Cleveland, OH 44106, USA

**Author notes:** **Corresponding Author** Daniel Dorfsman John P. Hussman Institute for Human Genomics 1501 NW 10^th^ Ave, Miami, FL 33136, USA.

## Abstract

**INTRODUCTION:** Cognitive SuperAgers (SA) are individuals aged 80+ with exceptional episodic memory performance for their age, exceeding middle-aged adult norms. This study integrates family- and association-based methods to identify genetic variants associated with SA in the Midwestern Amish population.

**METHODS:** 83 Amish SA were grouped into 16 pedigrees for parametric and non-parametric linkage analysis. Variants in linked regions (HLOD/LOD*≥3) were tested for association with SA using two contrasts: SA vs. Alzheimer’s disease (AD; n=40), and SA vs. cognitively unimpaired (CU), age-matched non-SA individuals (CU80+; n=157).

**RESULTS:** Evidence of linkage for SA was observed on chromosomes 1, 2, 7, 16, and 20, with the strongest signal around the AD-associated locus *WDR12* on chromosome 2. Association analysis for SA vs. AD identified eight variants in *HIVEP3* (chromosome 1) that were nominally significant when comparing SA vs. CU80+.

**DISCUSSION:** *WDR12* and *HIVEP3* are potential candidate genes contributing to SA in the Amish population.

## Background

The demographic shift towards older adults is expected to coincide with a rise in age-related conditions, underscoring the need for therapeutic advances.^1^ Cognitive health is a key determinant of well-being in later years, and cognitive decline that compromises independence is classified as dementia, with Alzheimer’s disease (AD) being the leading cause.^2^ The progressive nature of AD dementia—typically beginning with memory loss and advancing to impairment across multiple cognitive domains—often necessitates comprehensive care, making AD one of the costliest healthcare conditions.^2,3^

AD is heritable; genome-wide association studies (GWAS) have reported over 80 genetic loci linked to increased or decreased risk of late-onset AD (LOAD; symptom onset at ≥ 65 years) .^4^ The strongest of these genetic risk factors is the ε4 allele of the *APOE* gene, while most other AD-associated loci confer low-to-moderate effects.^5^ However, these discoveries largely result from studies using AD status as the primary outcome of interest. Given that the prevalence of AD increases markedly with advanced age, studying older adults with preserved or exceptional cognition may reveal protective genetic factors that delay or prevent disease.^2^

Studies of exceptional aging traits often employ specific phenotypic constructs to identify homogeneous groups of older adults. These range from basic longevity measures (e.g., centenarians) to composite phenotypes integrating age and additional measures of well-being, including physical or cognitive function.^6–9^ Given that episodic memory dysfunction is one of the most pronounced signs of AD, the cognitive SuperAger (SA) phenotype—defined as individuals age 80 and older with episodic memory performance exceeding that of a typical middle-aged adult—is a stark contrast to AD dementia, representing the opposite end of the memory spectrum.^10^ Therefore, studies of SA may be a strategy for identifying genetic factors that contribute to exceptional cognitive performance in older adults and those that protect against cognitive decline despite AD neuropathological changes.

The Midwestern U.S. Amish are a European-descended population known for their traditional lifestyle and endogamous practices. Originating from approximately 3,500 individuals who migrated to the U.S. in the 18^th^ and 19^th^ centuries, the Amish population has grown to over 400,000 by 2024.^11,12^ The Amish are widely regarded as an isolated founder group—a population derived from a bottleneck event, characterized by a closed gene pool. Founder populations like the Amish are strong candidates for genetic research on both Mendelian and complex traits, as their relative genetic homogeneity suggests traits are driven by fewer loci, facilitating the detection of phenotypic effects. Additionally, the availability of detailed Amish genealogical information enables the use of both genetic linkage- and association-based analytical frameworks, further enhancing the discovery potential within this population. Our group has previously leveraged these advantages to identify genetic loci associated with AD and Successful Aging—a broad phenotype encompassing preserved cognition and physical function.^13–17^ In the present study, we expand on this work by focusing on cognitive SuperAging, which is more narrowly defined and centers on exceptional episodic memory in late life, a domain particularly vulnerable to aging and AD neuropathology.^18^ By applying this targeted definition in the Amish, we aimed to identify genetic factors that 1) protect against AD and 2) promote exceptional memory performance into advanced age.

Our study objective was to identify genetic variants underlying exceptional episodic memory performance in older adults by conducting genetic linkage and association analyses in Amish with SA—a unique application of this phenotype in a genetic study. First, we identified Amish SA individuals using neuropsychological test criteria developed by the Northwestern University SuperAging Program.^10^ Utilizing genealogical data from the Anabaptist Genealogical Database (AGDB), we grouped these individuals into computationally tractable pedigrees for parametric and non-parametric linkage analysis.^19^ Genetic variants within linked regions were then tested for association using a case-control study design. We hypothesized that this approach would enhance our power to detect genetic effects and deepen understanding of both SuperAging and AD, potentially informing therapeutic strategies to delay or prevent AD.

## Methods

### Study Population

Participants in this study were enrolled through the Collaborative Amish Aging and Memory Project (CAAMP), a longitudinal study investigating cognitive and aging traits in the Midwestern Amish. CAAMP participants primarily reside in Adams, Elkhart, and LaGrange Counties in Indiana, and Holmes County in Ohio. Prior to recruitment, the research team consulted with Amish community leaders to develop an appropriate enrollment strategy; based on their recommendation, participation decisions were made at the individual level rather than through a community-level process. Recruitment initially prioritized individuals aged ≥ 80 with a cognitively impaired sibling. These criteria were later expanded to include all individuals ≥ 75.

Participants underwent a comprehensive neuropsychological test battery designed to assess performance across multiple cognitive domains. These include items from the Consortium to Establish a Registry for Alzheimer’s Disease (CERAD) battery, such as the Word List Learning, Category Fluency, Word List Recall, and Constructional Praxis tests.^20^ Additional measures include the Trail Making Tests (versions A and B) and the Multilingual Naming Test (MINT).^21,22^ All evaluations are reviewed by a clinical adjudication board comprising neurologists and psychologists, who apply established criteria to reach a consensus diagnosis of cognitively unimpaired (CU), AD dementia, mild cognitive impairment (MCI), or cognitively impaired not demented (CIND).^23–25^ Diagnoses of AD dementia follow the National Institute on Aging-Alzheimer’s Association (NIA-AA) clinical criteria, while CU status is assigned when cognitive performance falls within age-adjusted normative expectations across domains. SA are identified as CU individuals with episodic and non-episodic memory performance exceeding normative thresholds, as described in the following section. All diagnostic groups were drawn from the same CAAMP cohort and come from the same Amish kindreds within Indiana and Ohio.

### SuperAger Classification

#### Neuropsychological Tests Used to Define SuperAgers

The SA criteria developed by the Northwestern University SuperAging Program were applied in this study as closely as possible.^10,26,27^ In the original Northwestern University study, SA were defined as individuals aged ≥ 80 with episodic memory performance at or above the mean level of performance for individuals aged 56-64, along with non-episodic memory performance no lower than one standard deviation below the mean, for their age group.^26^ Episodic memory was assessed using the delayed recall component of the Rey Auditory Verbal Learning Test (RAVLT).^28^ Non-episodic memory tests included the Boston Naming Test, Trail Making Test B, and Category Fluency Test. ^29,30^ In CAAMP, the recall component of the Word List Learning test is most comparable to the RAVLT used in the original investigation, therefore it was used to assign SA status here. In addition, the Boston Naming Test has been replaced with the MINT in the National Alzheimer’s Coordinating Center (NACC) Uniform Data Set (UDS) to improve its compatibility across multiple languages; thus, the English version of the MINT was incorporated into the SA criteria for this study.^21,31^ The remaining tests in the original SA criteria—the Category Fluency and Trail Making Test B—were performed in CAAMP and were used as part of the SA classification.

#### SuperAger Test Performance Criteria

Age-, sex-, and education-specific normative values provided in version 3 of the NACC UDS were used for the Category Fluency, Trail Making Test B, and MINT components of the SA criteria.^31^ These values represent the cognitive performance of unimpaired UDS participants across various demographic subgroups. While the Amish typically receive 8 years of formal education, the UDS dataset’s most comparable education cutoff is :-s 12 years of education, therefore this threshold was applied. Since the normative values in the UDS primarily reflect older participants, an alternative source of normative information was required for the CERAD Word List Learning test. A cognitive screening that included the Word List Learning was administered to young-to-middle-aged participants in the third generation cohort of the Framingham Heart Study.^32^ A normative data report derived from this cohort provides an age-stratified performance summary for the following age groups: < 35, 35-44, 45-54, and ≥ 55. Initially, the mean performance for ages 45-54 (6.6 words recalled) was deemed the most relevant comparison group based on the 56-64 range established in the original Northwestern University SA definition. However, since recall scores are limited to integer values, the mean performance for the 35-44 age group (7 words recalled) was applied as the SA threshold. The Framingham normative values are reported as either age or education stratified, but not both simultaneously. Therefore, the cutoff of 7 in the 35-44 age group encompasses individuals with educational attainment ranging from :-s high school to graduate-level education.

#### Applying SuperAger Criteria to the Amish Sample

At the time of this study, 524 Amish participants had completed at least one evaluation using the relevant neuropsychological battery, received a consensus diagnosis, and had available genotype data. Of these, 92 individuals met SA criteria based on the normative thresholds. However, nine of these individuals received a consensus diagnosis of MCI or Unclear and were excluded from the analysis. This discordance between SA status and consensus diagnosis in these individuals may reflect subjective complaints, poor performance in cognitive domains outside the SA criteria, or other contextual factors contributing to a diagnosis other than CU.

### Genotyping and Imputation

Sample genotyping was performed on Illumina MEGAex+3k and GSA chips. The initial MEGAex+3k dataset comprised 2,038,867 variants genotyped in 1,343 individuals, while the GSA dataset contained 708,956 variants genotyped in 1,881 individuals. Both sets were processed using a common quality control (QC) procedure, which applied sample-level filters for genotype missingness (≥ 3%), duplicated samples, and sex mismatches. At the variant level, single nucleotide polymorphisms (SNPs) that were monomorphic or exhibited high levels of missingness (≥ 3%) were excluded. After this initial QC process, the MEGAex+3k dataset retained 959,578 variants across 1,298 individuals, and the GSA dataset retained 549,704 variants across 1,747 individuals. The two datasets were subsequently merged, retaining 329,071 common SNPs. The merged dataset underwent a similar QC procedure, including sample-level filtering to remove flagged duplicates and Mendelian errors, and variant-level filtering to remove SNPs with significant Hardy-Weinberg Equilibrium deviations (HWE; p < 1× 10^-6^). The final merged dataset comprised 329,055 variants across 2,973 individuals.

The combined dataset was imputed using the TOPMed imputation server with the TOPMed reference panel (version R2), which includes reference haplotypes from Pennsylvania Amish individuals.^33^ Haplotype phasing was performed with the Eagle algorithm.^34^ Following imputation, variant-level QC was applied as follows: rare variants (MAF < 1%) with an imputation quality (R^2^) < 0.8, common variants with R^2^ < 0.6, and multiallelic variants were excluded. The final imputed dataset contained 2,973 samples and genotype doses at 12,570,541 SNPs.

### Linkage Analysis

To assess the evidence of genetic linkage to the SA phenotype, SA individuals were subdivided into a set of computationally manageable pedigrees using an all-connecting-paths (ACP) pedigree generated from relationship data maintained in the AGDB.^19^ At the time of this study, all Amish participants in CAAMP could be connected in a single multigeneration pedigree comprising 12,659 individuals. The ACP pedigree was subdivided using PedCut, which algorithmically groups subjects of interest (SOI) into the smallest possible number of subpedigrees, each containing the maximum number of SOI connected by a common ancestor.^35^ Using the ACP, the software grouped the 83 SA individuals into 16 pedigrees, as shown in Supplementary Figure 1.

The SA subpedigrees were used to prepare a linkage dataset in PLINK format.^36^ Genotyped autosomal markers were selected from the pre-imputation QC dataset. A subset of variants in low linkage disequilibrium (LD) were selected by filtering for minor allele frequency (MAF) > 30%, then pruning using a 500 kb window size, 1-marker step size, and r^2^ threshold of 0.05. The resulting dataset contained 7,559 autosomal markers. Genetic map positions were obtained from the Illumina GSA support files, and are derived from the deCODE recombination maps.^37^

Multipoint, affecteds-only linkage analysis was performed in MERLIN using both parametric (dominant and recessive) and nonparametric models.^38^ Parametric analyses calculated heterogeneity LOD scores (HLOD) to account for potential locus heterogeneity. In the dominant parametric linkage analysis, the trait allele frequency was set to 0.05, and the penetrance values were set to 0, 0.95, and 0.95 for non-carriers, heterozygotes, and homozygote carriers, respectively. For the recessive parametric linkage analysis, the trait allele frequency was set to 0.2, and the penetrance values were set to 0, 0, and 0.95 for non-carriers, heterozygotes, and homozygote carriers, respectively. Non-parametric linkage analysis calculated Kong and Cox LOD* scores using the Whittemore and Halpern S_all_ scoring function, which considers allele sharing across all SA individuals in the pedigree.^39,40^ HLOD or LOD* scores exceeding 3 were considered evidence of significant linkage warranting follow-up. Variants within a 1-LOD-down support interval—defined as the genomic region surrounding the linkage peak where the score decreases by one unit—were prioritized for genetic association analysis.

### Genetic Association Testing

Tests of association between the SA phenotype and genetic variants were performed using the R/Bioconductor GENESIS package.^41^ A generalized linear mixed model framework was utilized to account for the increased kinship within the Amish sample. SNPs with a MAF > 1%, located within a 1-LOD-down support interval of linked regions of interest were analyzed. Principal components (PCs) and pairwise kinship estimates were generated using the GENESIS PC-AiR and PC-Relate methods, respectively, utilizing an LD-pruned (r^2^ < 0.16), genome-wide panel of genotyped SNPs.^41^

Association analyses utilized an outcome of SA, and comparison groups comprising either AD or CU aged ≥ 80 individuals (CU 80+). The rationale for these comparisons is twofold: 1) comparing AD to SA employs an “extreme phenotype” sampling approach, potentially enhancing the detection of associations by minimizing phenotypic heterogeneity in both groups; and 2) genetic associations identified when comparing SA to age-matched CU may indicate the presence of variants that contribute to exceptional cognitive performance in unimpaired older adults. The null model in these analyses was adjusted for the fixed effects of age, sex, and PCs 1 and 2, and a random effect kinship matrix. Members of the Amish community—including those enrolled in CAAMP—largely receive eight years of formal education; therefore, due to minimal variability within the sample, education was omitted as a covariate. To establish an adjusted Bonferroni threshold for significance in each region (0.05/number of independent tests), the SimpleM procedure was applied to estimate the number of independent tests performed.^42^ In addition, given the disproportionately strong effect of the *APOE* ε4 allele, associated SNPs were reexamined in a secondary model that included ε4 carrier status as an additional covariate to assess whether associations were independent of *APOE*-related risk.

## Results

### SuperAger demographics and neuropsychological performance compared to AD and CU 80+

Age, sex, *APOE* ε4 carrier frequency, and SA-defining neuropsychological test performance for SA (n=83), CU 80+ (n=157), and AD (n=40) participants are presented in Table 1. SA individuals demonstrated significantly better performance on all neuropsychological tests compared to those with AD. The SA group was, on average, significantly younger than the AD group (82.9 versus 85.4 years, p = 4.7 × 10^-3^). While the proportion of women was higher in the SA group compared to the AD group (69% versus 60%), this difference was not statistically significant. Not surprisingly, the frequency of the *APOE* ε4 allele was significantly lower among SA compared to AD (18% versus 60%, p = 2.9 × 10^-6^).

**Table 1.**
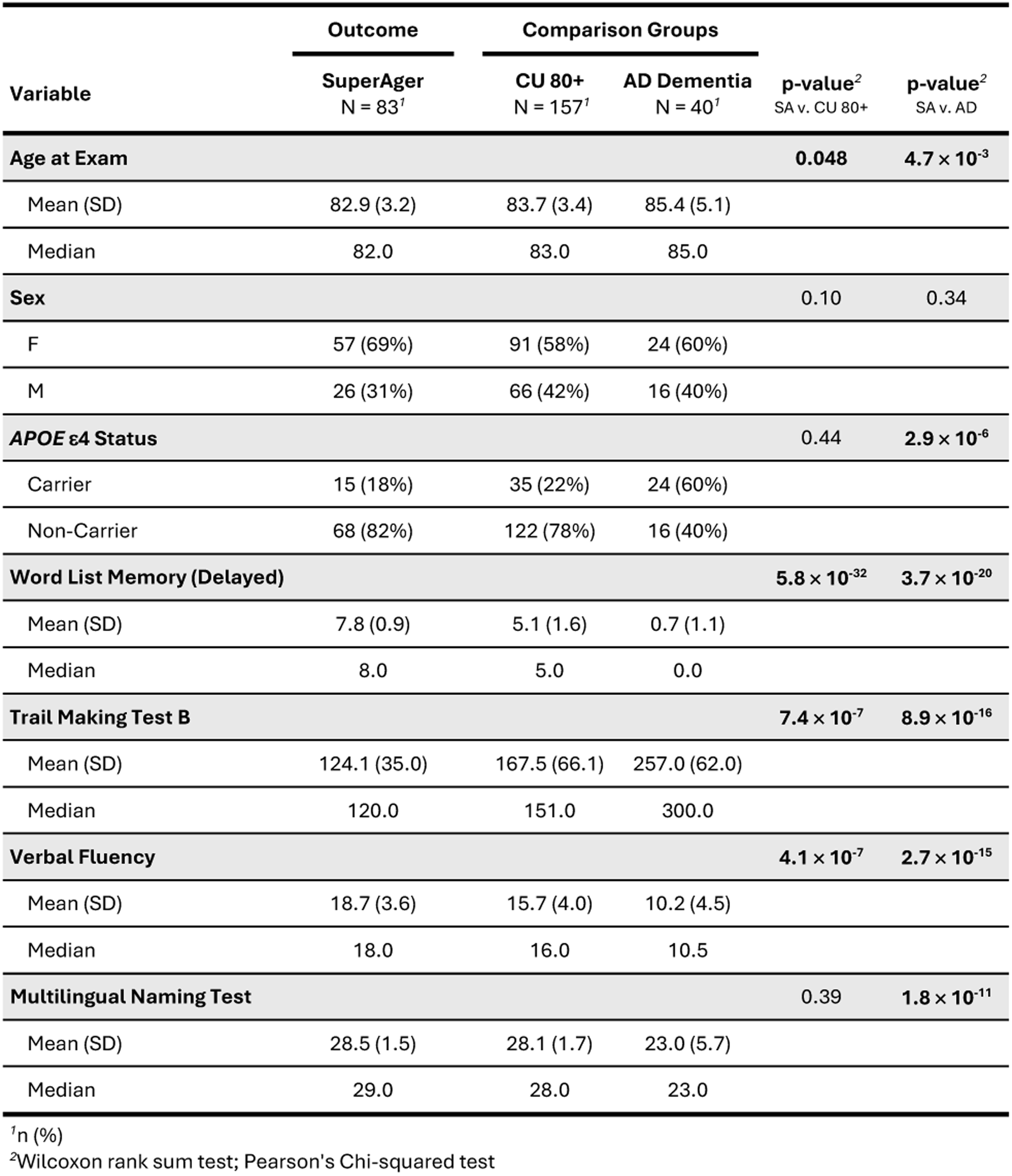
Summary of age, sex, APOE ε4 carrier status, and SuperAger-defining neuropsychological test performance for SuperAgers, CU 80+, and individuals with AD dementia.

When compared to CU 80+, SA individuals performed significantly better on the Word List Memory, Trail Making Test B, and Verbal Fluency tests, but no significant differences were observed on the Multilingual Naming Test. There was a small but significant difference in age between SA and CU 80+ (82.9 versus 83.7 years, p = 0.048). Women were similarly overrepresented in the SA group compared to CU 80+ (69% versus 58%), though this difference was not statistically significant. There was a smaller—though non-significant—proportion of *APOE* ε4 carriers among SA than CU 80+ (18% versus 22%).

### Linkage Results

The genome-wide multipoint parametric (dominant and recessive) and nonparametric linkage results are presented in Supplementary Figure 2. In the dominant parametric analysis, HLOD scores exceeding 3 were identified on chromosomes 1 (peak at 44.7 Mb, HLOD =3.10), 2 (202.9 Mb, HLOD = 3.92), 7 (30.2 Mb, HLOD = 3.14), and 20 (16.7 Mb, HLOD = 3.71). In the recessive parametric analysis, HLOD scores greater than 3 were observed on chromosomes 2 (205.2 Mb, HLOD = 3.03), 16 (22.7 Mb, HLOD = 3.18), and 20 (16.7 Mb, HLOD = 3.17). In the nonparametric analysis, a LOD* score greater than 3 was observed on chromosome 2 (204.2 Mb, LOD* = 3.20). The peak results from both parametric analyses, in addition to the non-parametric analysis, are summarized in Table 2.

**Table 2.** Summary of results for parametric and non-parametric linkage analysis in SuperAger pedigrees.

| Position<sup>1</sup> | SNP ID | LOD | HLOD | LOD* | Function | Nearest Gene | Model |
| --- | --- | --- | --- | --- | --- | --- | --- |
| **Chr 1** |  |  |  |  |  |  |  |
| 44667889 | rs6683133 | 2.84 | 3.10 | - | Intronic | *TMEM53* | Dominant |
| **Chr 2** |  |  |  |  |  |  |  |
| 202875949 | rs3845802 | 2.46 | 3.92 | - | UTR3 | *WDR12* | Dominant |
| 205197825 | rs1921788 | 1.74 | 3.03 | - | Intronic | *PARD3B* | Recessive |
| 204481944 | rs12613305 | - | - | 3.20 | Intergenic | *ICOS;PARD3B* | Non-Parametric |
| **Chr 7** |  |  |  |  |  |  |  |
| 30155442 | rs7780166 | 1.42 | 3.14 | - | Intronic | *MTURN* | Dominant |
| **Chr 16** |  |  |  |  |  |  |  |
| 22707617 | rs154533 | 2.86 | 3.18 | - | Intergenic | *OTOAP1;HS3ST2* | Recessive |
| **Chr 20** |  |  |  |  |  |  |  |
| 16685245 | rs6034572 | 2.90 | 3.71 | - | Intergenic | *KIF16B;SNRPB2* | Dominant |
| 16685245 | rs6034572 | 2.33 | 3.17 | - | Intergenic | *KIF16B;SNRPB2* | Recessive |

<sup>1</sup>Build hg38.

### Regional Association Analysis

#### Follow-Up of Known AD Locus *WDR12*

Cross-referencing the linkage peaks with known AD loci revealed that the strongest overall signal, observed on chromosome 2 during the parametric dominant analysis (rs3845802, HLOD_dominant_ = 3.92), colocalized with a recently reported AD risk locus in *WDR12*.^4^ A GWAS conducted by Bellenguez et al. (2022) identified a small insertion/deletion polymorphism (indel) at rs139643391 in *WDR12* (minor allele = T, major allele = TC), with the minor allele associated with reduced odds of AD compared to unaffected controls (OR = 0.94).^4^ This locus has since been incorporated into the Alzheimer’s Disease Sequencing Project (ADSP) list of AD loci with genetic evidence.^43^ The recessive and non-parametric analyses, while exhibiting their maximum linkage peaks upstream of the dominant signal, also demonstrated modest evidence of linkage at the peak dominant marker rs3845802 (HLOD_recessive_ = 2.82, LOD* = 3.19).

To further investigate this result, the established *WDR12* variant rs139643391 was tested for association with SA in the Amish. While the rs139643391 variant was absent from the QC’d imputed dataset, the genomic boundaries of *WDR12* as defined by Ensembl—encompassing the union of its alternative transcripts—contained 129 SNPs in complete LD (r^2^ = 1) with rs139643391.^44^ Tests of association between SA and AD revealed these SNPs to be nominally associated with increased odds of SA (p = 0.035, OR = 3.1). However, no associations were observed when comparing SA to CU 80+ (p = 0.93, OR = 0.97). Combining the SA and CU 80+ and evaluating the association of the 129 *WDR12* variants using AD as the comparison group found increased odds of being cognitively unaffected (SA and CU 80+, combined) versus affected with AD (p = 0.018, OR = 2.55). A highlighted subset of the 129 tests, corresponding to SNPs resulting in nonsynonymous amino acid substitutions, are shown in Supplementary Table 1. After adjustment for *APOE* ε4, the association strengthened modestly (OR = 4.2, p = 0.17) in the SA versus AD comparison, consistent with an effect that is independent of *APOE*-related risk. The *APOE*-adjusted results for functionally relevant SNPs in the *WDR12* locus are shown in Supplementary Table 2.

Examination of the SA subpedigrees (Supplementary Figure 1) for the protective allele in *WDR12* revealed notable enrichment in subpedigree 6, where five of the six SA members carried the protective allele, and in subpedigree 7, where three of the five members were carriers. These two subpedigrees also showed the highest family-specific LOD scores in the dominant parametric analysis (LOD_subpedigree_6_ = 2.3; LOD_subpedigree_7_ = 1.02). Tracing the eight *WDR12* protective allele carriers from subpedigrees 6 and 7 through the ACP pedigree identified a most recent common ancestor eight generations back, suggesting that the protective allele is segregating broadly within the Amish population.

#### Regional Association in 1-LOD Support Intervals

A summary of the 1-LOD support intervals on chromosomes 1, 2, 7, 16, and 20, including the total number of variants tested in each region and the SimpleM adjusted significance thresholds, is provided in Supplementary Table 3. For chromosomes 2 and 20, an HLOD* or LOD* greater than 3 was observed across multiple analyses (e.g., both dominant and recessive models). In these cases, the 1-LOD interval applied during the association analyses was calculated based on the strongest linkage peak, which, in both instances, was derived from the parametric dominant analysis.

When comparing SA to AD, intronic SNP rs12734651, located in the *HIVEP3* gene on chromosome 1, was significantly associated with SA (OR = 0.24, p = 6.46 × 10^-6^), with the minor allele (A; MAF_CAAMP_ = 30%) overrepresented among individuals with AD. This SNP was strongly correlated with seven additional intronic SNPs that were also significantly associated, as shown in Figure 1. The top SA-AD results for each chromosome are presented in Supplementary Table 4. When comparing SA individuals to CU 80+, no associations exceeding the adjusted thresholds were observed. The SA-CU 80+ results are summarized in Supplementary Table 5.

**Figure 1.**
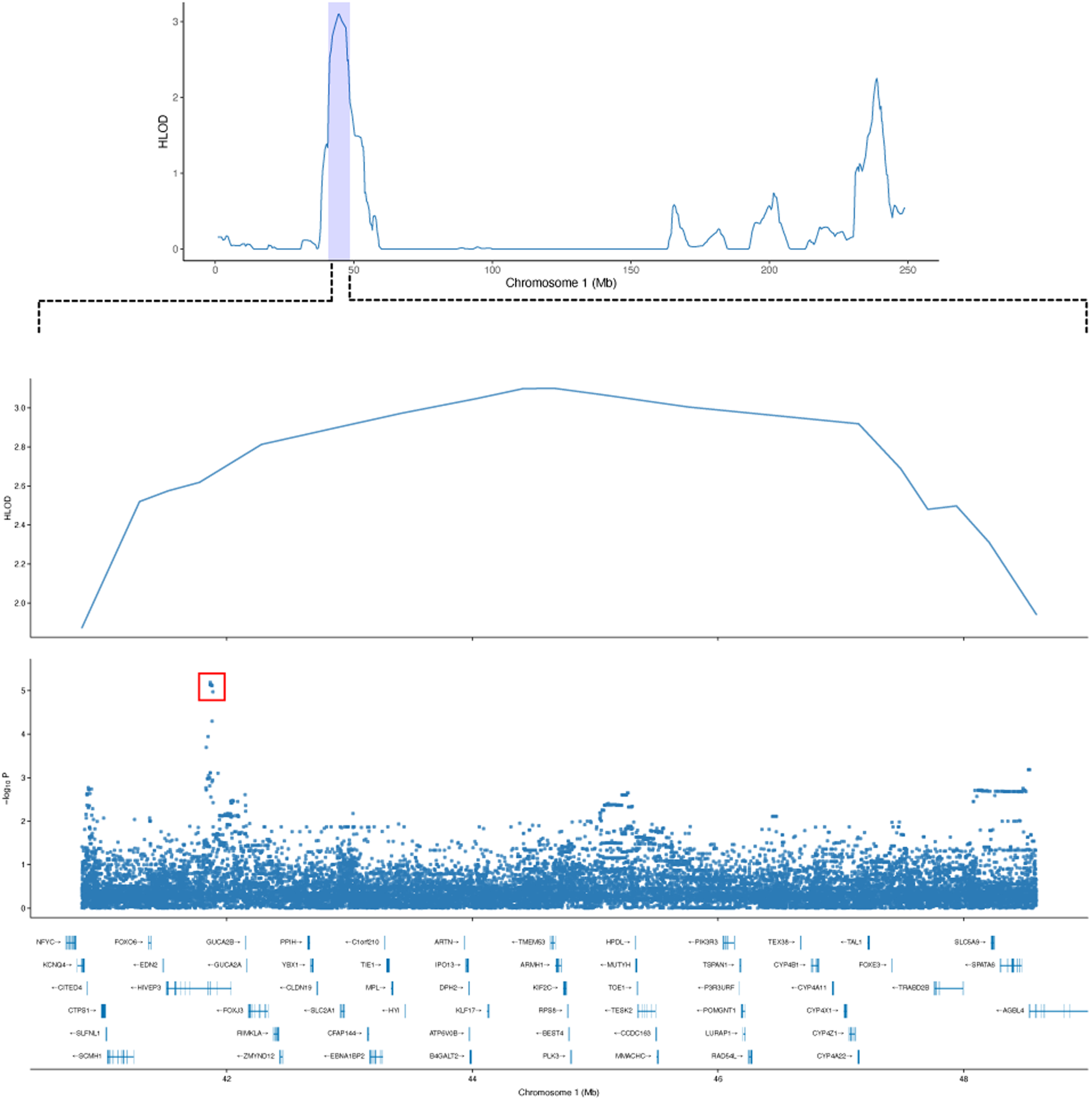
**Significant association of SNPs in the chromosome 1 linkage region with the SuperAger phenotype, relative to AD** The SA dominant parametric linkage plot for chromosome 1, highlighting the 1-LOD support interval, is shown at the top of the figure. The adjacent panels underneath illustrate the peak linkage region, along with the single-variant results from the SA-AD analysis. The strongest association is detected within the HIVEP3 gene (indicated by the red box).

To compare the associations observed in *HIVEP3* between the SA-AD and SA-CU 80+ analyses, we selected the eight *HIVEP3* variants identified in the SA-AD analysis. Notably, these variants were also nominally associated with SA compared to CU 80+ (OR = 0.61, p = 0.032). Similarly, the minor allele (A) at the peak SNP rs12734651 was overrepresented in CU 80+ relative to those in the SA group. Summary statistics for all eight variants across both comparisons are presented in Supplementary Table 6. Effect size estimates for *HIVEP3* variants were largely unchanged following *APOE* adjustment, although p-values were modestly attenuated in the *APOE*-adjusted model when comparing SA to AD (rs12734651; p = 1.99 × 10^-4^, OR = 0.26), likely due to the inclusion of additional variables in the model. Summary statistics for the *APOE*-adjusted *HIVEP3* results are shown in Supplementary Table 7.

## Discussion

This study aimed to explore the SuperAger phenotype in the Amish population to identify genetic factors associated with exceptional cognitive function in older adults. By integrating family- and population-based analytical approaches, multiple loci of interest were identified, including *WDR12 and HIVEP3*, which may influence the SuperAger phenotype in this population. Notably, *WDR12* has recently been linked to AD risk.^4^ Emerging evidence also suggests a potential involvement of *HIVEP3* in AD-related characteristics such as hippocampal volume and cognitive trajectories.^45–47^ Taken together, these findings suggest that these loci represent plausible genetic factors underlying both AD and the SuperAger phenotype, and that variants associated with exceptional episodic memory after 80 years of age might be responsible for some of the decreased risk of AD conferred by these loci.

The SuperAger linkage analysis identified regions of interest on chromosomes 1, 2, 7, 16, and 20. The strongest signal was found within the recently reported AD locus *WDR12*, a ubiquitously expressed gene involved in ribosomal biogenesis that has also been implicated in cardiovascular outcomes, including coronary artery disease and myocardial infarction risk.^48–50^ This finding was particularly noteworthy given that a variant in *WDR12* (rs139643391) was associated with AD in the recent GWAS by Bellenguez et al.^4^ While the reported effect of this variant was modest (OR = 0.94), it represents a relatively low-frequency allele (MAF_Bellenguez et al._ = 13.1%; MAF_CAAMP_ = 13.6%) associated with reduced odds of AD, thus categorizing it as a “protective” allele. The single-variant association analysis in the present study confirmed the protective effect—through variants in complete LD with rs139643391—at a nominal significance threshold (α < 0.05), using SuperAgers as the outcome and AD as the comparison group (p = 0.035, OR = 3.1). Specifically, the frequency of the “protective” T allele was 5% in the AD group (present in 4 out of the 40 individuals with AD) compared to 13.3% among SuperAgers (present in 21 out of the 83 SuperAgers). However, expanding the analysis to include all cognitively unimpaired individuals over age 80 (CU 80+), the frequency of this allele was 13.6% in this group, which was not significantly different compared to its frequency in SuperAgers. This finding suggests that the protective allele may represent a general marker associated with cognitive preservation in older adults, rather than being uniquely enriched in SuperAgers. Nevertheless, the observation that subpedigrees 6 and 7, which drove the dominant parametric linkage signal, were enriched for the established protective *WDR12* allele strongly suggests that this allele—or a functionally relevant allele in LD—underlies the overall linkage signal. The marginal evidence of linkage from the remaining subpedigrees could reflect allelic heterogeneity, where multiple distinct variants in the region influence the phenotype through distinct mechanisms. Furthermore, the colocalization of the aggregate linkage signal to this specific region, which has been implicated as a potential modulator of AD risk, reinforces its relevance to cognitive phenotypes, including SuperAging.

The regional association analysis in other linkage regions identified eight SNPs (peak signal at rs12734651) associated with SuperAging compared to AD in the *HIVEP3* gene on chromosome 1. The estimated effect of the associated variant suggests that the minor allele increases the odds of AD relative to being a SuperAger. These variants were also nominally associated with the SuperAger phenotype when compared to CU 80+. Similarly, the less common allele was estimated to increase the odds of CU 80+ compared to SuperAgers. Together these findings suggest that the minor allele may: 1) increase the risk of AD, 2) increase the likelihood of CU 80+ relative to being SuperAger. This observation is notable because it indicates that variants in *HIVEP3* may influence AD and contribute to variability in cognitive performance among cognitively unimpaired older adults. The common allele (C)—associated with increased odds of being a SuperAger—had a frequency of 80% among SuperAgers, 48.8% among those with AD, and 71% among CU 80+. Of the 83 SuperAgers, 79 carried at least one copy of the C allele in rs12734651.

The eight associated SNPs lie within a 15 kilobase intronic region of *HIVEP3*. Examining this interval in the UCSC Genome Browser indicates that these SNPs fall within regions identified by GeneHancer as putative enhancers, suggesting they are located in regulatory blocks that may modulate *HIVEP3* expression, as shown in Supplementary Figure 3.^51,52^ If these variants influence transcription factor binding or chromatin accessibility, they could alter enhancer activity and impact *HIVEP3* regulation.

*HIVEP3* is a member of the *HIVEP* gene family, which encodes zinc finger proteins and includes *HIVEP1*, *HIVEP2*, and *HIVEP3*. The *HIVEP3* gene exhibits biased expression in the brain and, through its interaction with the kB binding motif, has been implicated as a modulator of inflammatory and apoptotic pathways.^53,54^ Notably, neuroinflammation is now widely recognized as a key driver of AD-related neuropathological changes.^55^ An earlier study reported evidence of association between variants in *HIVEP3* and Parkinson disease.^56^ More recently, variants in *HIVEP3* have been associated with increased amyloid deposition and reduced hippocampal volume in cognitively unimpaired older adults, as well as an increased overall risk of AD.^45,46^ In addition, a GWAS performed in a cohort of amyloid-positive older adults without cognitive impairment identified a genome-wide significant variant in *HIVEP3* associated with differences in cognitive trajectory.^47^ Supporting these genetic findings, a recent transcriptomic analysis of synaptosomes in post-mortem brain tissue identified *HIVEP3* among the top upregulated genes in AD versus control synapses.^57^ The variants identified in these prior studies are distinct and uncorrelated (based on R^2^) with those reported here, suggesting that allelic heterogeneity at this locus may confer related outcomes. The present study’s findings—indicating that *HIVEP3* may influence both AD risk and cognitive performance in unimpaired individuals—together with prior evidence linking variants in *HIVEP3* to AD-related mechanisms, support its role as a candidate locus for both AD and SuperAging.

This study has certain limitations that warrant consideration. The unique features of the Amish population (e.g., years of education, occupation) pose challenges when applying normative data derived from non-Amish sources, potentially introducing classification error. The normative data used to identify Amish SuperAgers were derived from non-Amish populations, which may differ from the average performance of the Amish on these tests. Furthermore, the Rey Auditory Verbal Learning Test (RAVLT), used to measure episodic memory in the original SuperAger study, is a general-purpose test of memory performance. In contrast, the CERAD Word List Learning test used in this investigation is specifically designed for older adults and is less complex. While the normative threshold for identifying SuperAgers among the Amish was based on the average performance of unimpaired middle-aged adults, it may have lower specificity than the RAVLT for identifying individuals with truly exceptional memory performance. Lastly, the findings reported here are interpreted in the context of predominantly non-Hispanic European ancestry populations, both in the CAAMP cohort and in the supporting studies used for comparison. Thus, the generalizability of these associations to other ancestral populations is not yet fully established.

In summary, these findings highlight *WDR12* and *HIVEP3* as potential loci influencing the SuperAger phenotype in the Amish population. The observed linkage to *WDR12* was driven by a subset of SuperAger pedigrees carrying a known protective allele for AD at this locus, while the evidence for *HIVEP3* indicates the enrichment of common trait-associated alleles among SuperAgers compared to both AD and CU 80+ individuals, suggesting promotion of exceptional memory in these individuals, rather than simply preventing progression of AD in individuals with normal memory for their ages. Importantly, leveraging Amish pedigree information enabled the identification of loci that may have been overlooked in traditional study designs, supporting the value of continued genetic research in this unique population. Together, these findings underscore the complex genetic architecture underlying both AD risk and the SuperAger phenotype and suggest that *WDR12* and *HIVEP3* are strong candidates for functional genomic characterization to reveal mechanisms that promote superior cognitive functioning in older adults and guide the development of AD interventions.

## Data Availability

The data referenced in this manuscript is available upon request and with principal investigator approval by reaching out to the primary author.

