## Supplementary Material for "*WDR12* and *HIVEP3* are Contributors to Cognitive Preservation in Amish SuperAgers"

**Supplementary Figure 1. Subpedigrees (n=16), derived from an ACP pedigree generated by the AGDB, encompassing all 83 Amish SuperAgers (sex-neutral symbols used to protect participant privacy)**

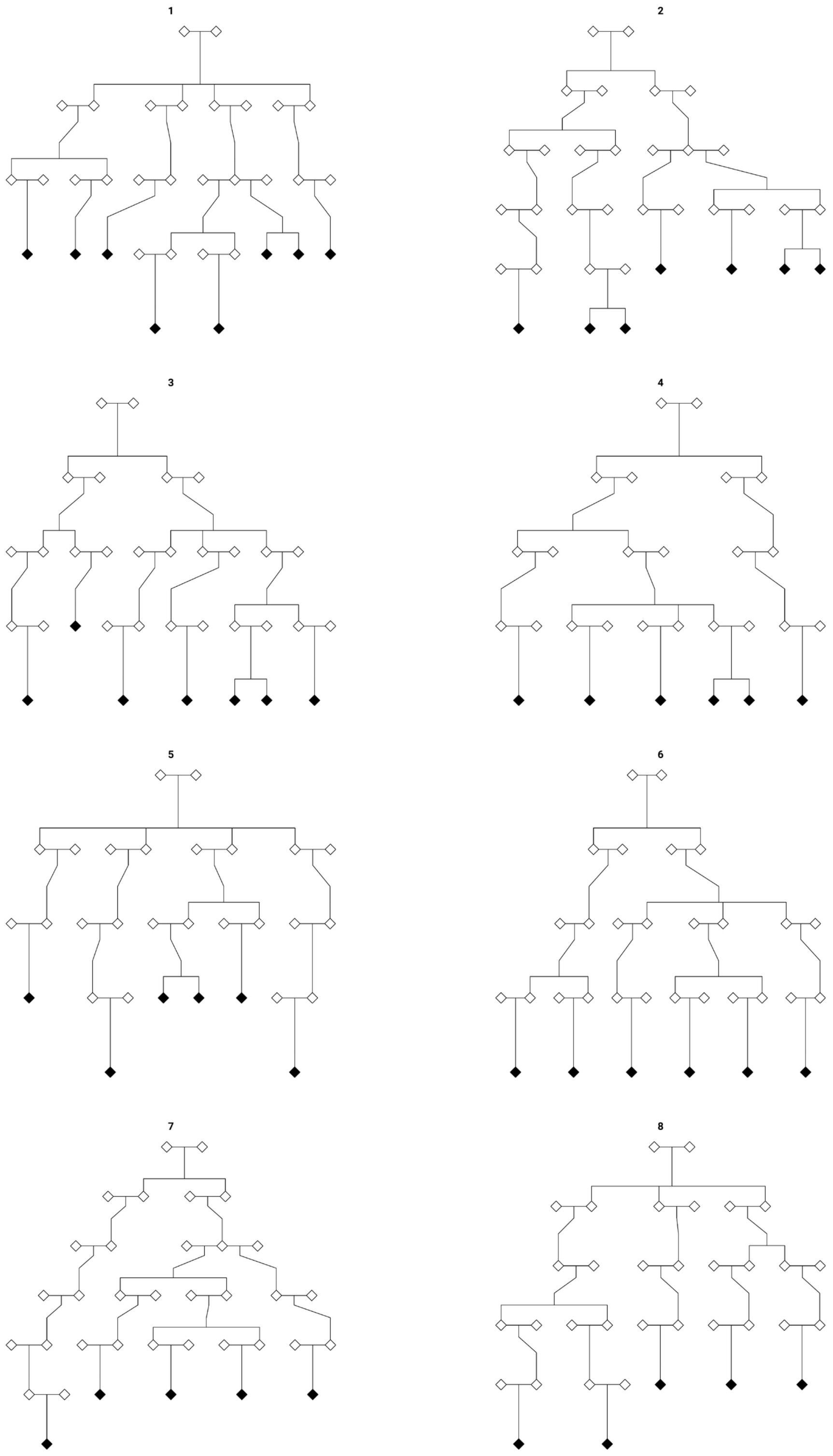

**9**

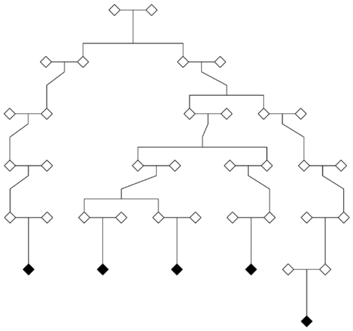

**10**

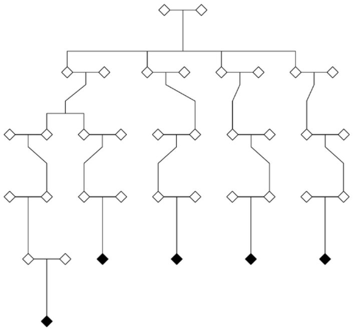

**11**

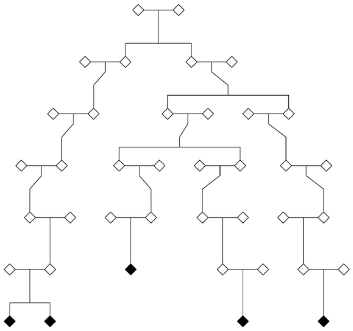

**12**

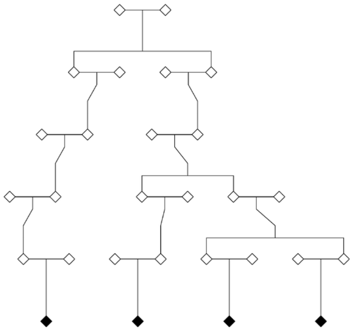

**13**

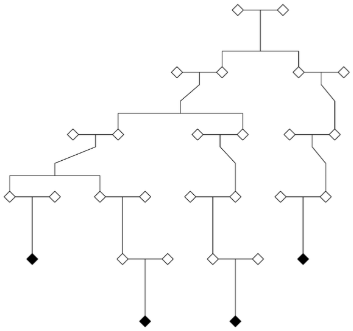

**14**

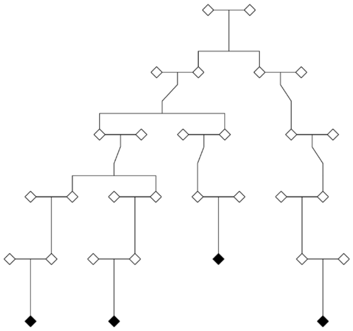

**15**

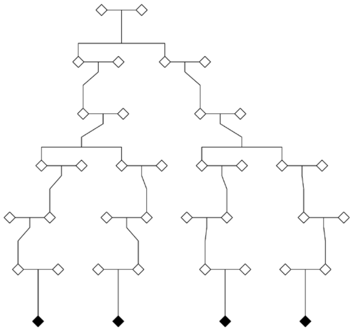

**16**

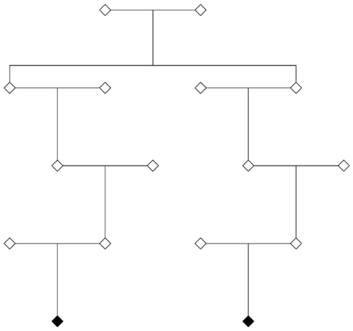

**Supplementary Figure 2. SuperAger genome-wide linkage plots**

The genome-wide linkage plots below show the results of the parametric dominant, parametric recessive, and non-parametric linkage analyses. A linkage threshold of HLOD or LOD* > 3 was used for regional follow-up, indicated by the dotted red lines in each sub-plot. The dominant parametric identified HLOD scores > 3 on chromosomes 1 (peak at 44.7 Mb, HLOD = 3.1), 2 (202.9 Mb, HLOD = 3.9), 7 (30.2 Mb, HLOD = 3.1), and 20 (16.7 Mb, HLOD = 3.7). In the recessive parametric analysis, HLOD scores > 3 were observed on chromosomes 2 (205.2 Mb, HLOD = 3.0), 16 (22.7, HLOD = 3.2), and 20 (16.7 Mb, HLOD = 3.2). In the non-parametric analysis, a LOD* score greater than 3 was observed on chromosome 2 (204.2 Mb, LOD* = 3.2).

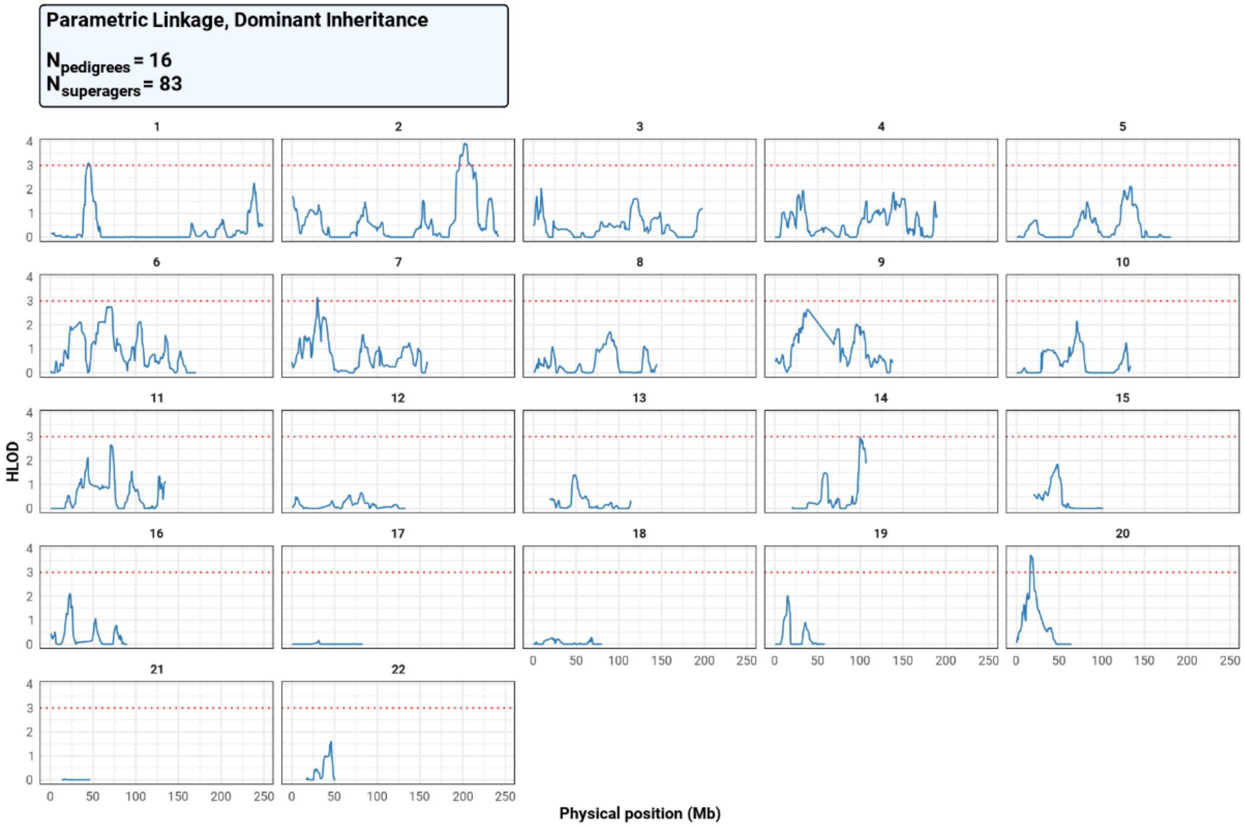

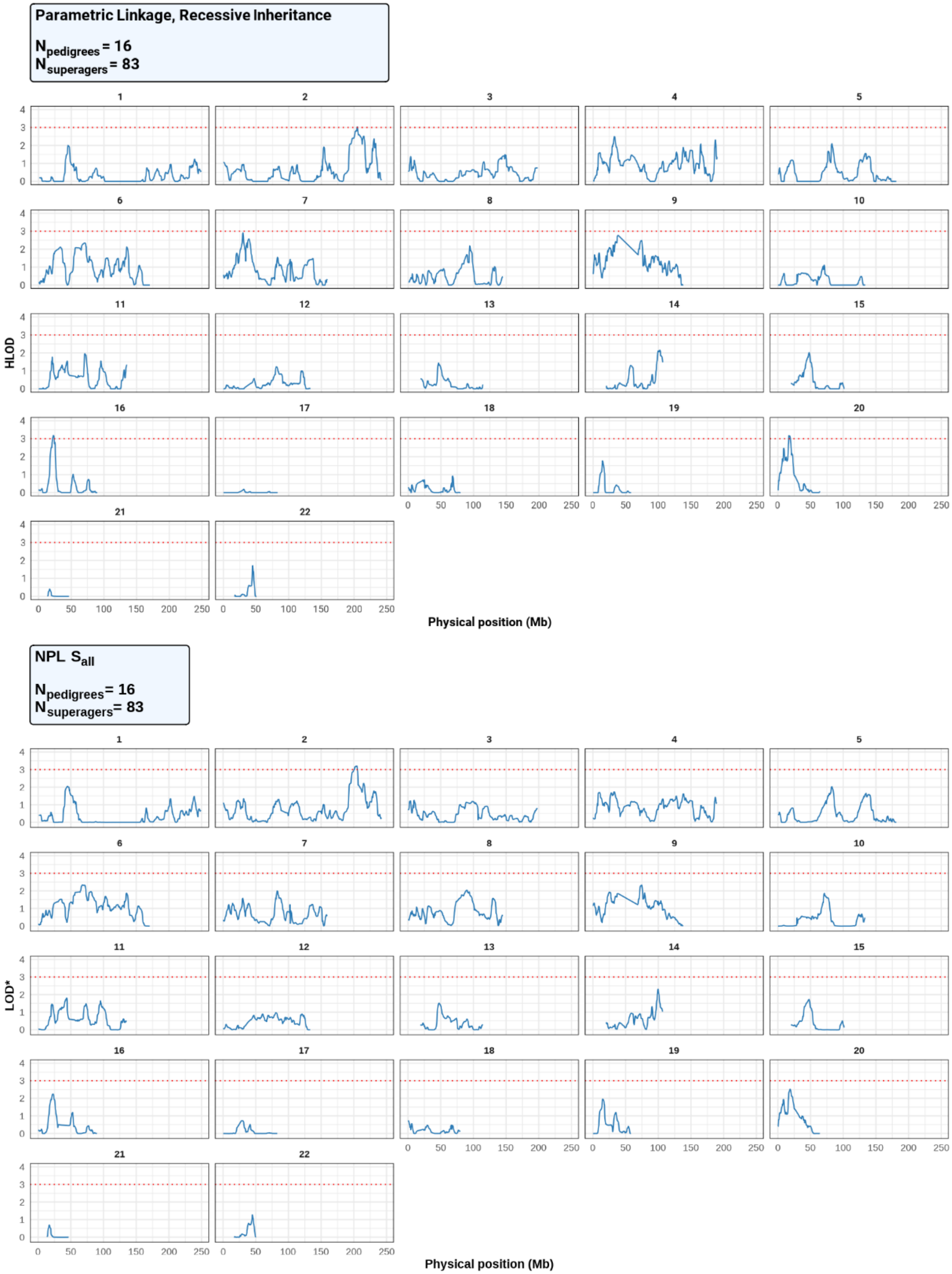
Parametric Linkage, Recessive Inheritance
$N_{pedigrees}$ = 16
$N_{superagers}$ = 83
HLOD
Physical position (Mb)
NPL $S_{all}$
$N_{pedigrees}$ = 16
$N_{superagers}$ = 83
LOD*
Physical position (Mb)

### Supplementary Table 1. Functionally relevant single-variant association results in *WDR12* region

| | | | | SuperAger vs. AD | | SuperAger vs. CU 80+ | | (SuperAger and CU 80+) vs. AD | | | | |
|---|---|---|---|---|---|---|---|---|---|---|---|---|
| Chr | Position[1] | ID | MAF | OR | P | OR | P | OR | P | Exonic Function | Gene | CADD Score[2] |
| 2 | 202901033 | rs35212307 | 0.14 | 3.1 | 0.035 | 0.97 | 0.93 | 2.6 | 0.018 | Nonsynonymous | *WDR12* | 22.4 |
| 2 | 202982094 | rs72932557 | 0.14 | 3.1 | 0.035 | 0.97 | 0.93 | 2.6 | 0.018 | Nonsynonymous | *CARF* | 15.8 |

[1]Build hg38.

[2]Phred-scale score: values > 20 indicate the top 1% of variants predicted to significantly alter function.

### Supplementary Table 2. Functionally relevant single-variant association results in *WDR12* region (adjusted for *APOE* ε4 carrier status)

| | | | | SuperAger vs. AD | | SuperAger vs. CU 80+ | | (SuperAger and CU 80+) vs. AD | | | | |
|---|---|---|---|---|---|---|---|---|---|---|---|---|
| Chr | Position[1] | ID | MAF | OR | P | OR | P | OR | P | Exonic Function | Gene | CADD Score[2] |
| 2 | 202901033 | rs35212307 | 0.14 | 4.2 | 0.017 | 0.96 | 0.90 | 2.9 | 0.012 | Nonsynonymous | *WDR12* | 22.4 |
| 2 | 202982094 | rs72932557 | 0.14 | 4.2 | 0.017 | 0.96 | 0.90 | 2.9 | 0.012 | Nonsynonymous | *CARF* | 15.8 |

[1]Build hg38.

[2]Phred-scale score: values > 20 indicate the top 1% of variants predicted to significantly alter function.

**Supplementary Table 3. Summary of genomic intervals, total loci, and independent tests for 1-LOD regions analyzed for association following SuperAger linkage analysis**

| Chromosome | Start Position[1] | End Position[1] | Total Loci | Independent Tests[2] | Adjusted Alpha[3] |
|---|---|---|---|---|---|
| 1 | 40820951 | 48591023 | 19386 | 4552 | $1.10 \times 10^{-5}$ |
| 2 | 195510796 | 211510661 | 39225 | 9232 | $5.42 \times 10^{-6}$ |
| 7 | 27423751 | 32811906 | 17499 | 4096 | $1.22 \times 10^{-5}$ |
| 16 | 18000830 | 25765598 | 19245 | 4800 | $1.04 \times 10^{-5}$ |
| 20 | 15650264 | 20826316 | 17761 | 4152 | $1.20 \times 10^{-5}$ |

[1]Build hg38.

[2]Independent Tests based on SimpleM calculation.

[3]Adjusted Alpha = 0.05 / Independent Tests

**Supplementary Table 4. Summary of results from single variant association tests comparing SuperAgers to individuals with AD**

| Chromosome | ID | Position[1] | MAF | OR | P | Function | Nearest Gene |
|---|---|---|---|---|---|---|---|
| 1 | rs12734651 | 41864914 | 0.30 | 0.24 | **$6.46 \times 10^{-6}$** | intronic | *HIVEP3* |
| 2 | rs11893875 | 200450045 | 0.11 | 0.17 | $3.24 \times 10^{-4}$ | intronic | *SPATS2L* |
| 7 | rs10486507 | 32360221 | 0.20 | 0.20 | $3.50 \times 10^{-5}$ | intronic | *PDE1C* |
| 16 | rs75910943 | 22964576 | 0.057 | 0.11 | $3.79 \times 10^{-4}$ | intergenic | *HS3ST2;USP31* |
| 20 | rs6080245 | 16366945 | 0.11 | 6.3 | $7.04 \times 10^{-4}$ | UTR3 | *KIF16B* |

[1]Build hg38.
Cells highlighted in bold indicate statistically significant results relative to the SimpleM adjusted threshold

**Supplementary Table 5. Summary of results from single variant association tests comparing SuperAgers to CU 80+ individuals**

| Chromosome | ID | Position[1] | MAF | OR | P | Function | Nearest Gene |
|---|---|---|---|---|---|---|---|
| 1 | rs76825575 | 42327990 | 0.038 | 6.9 | $3.23 \times 10^{-4}$ | intronic | *FOXJ3* |
| 2 | rs16850280 | 196362647 | 0.058 | 4.6 | $8.47 \times 10^{-4}$ | intronic | *HECW2* |
| 7 | rs17156996 | 28667310 | 0.011 | 50 | $7.41 \times 10^{-4}$ | intronic | *CREB5* |
| 16 | rs71375619 | 24494863 | 0.011 | 62 | $9.48 \times 10^{-5}$ | intergenic | *CACNG3;RBBP6* |
| 20 | rs112640297 | 16304818 | 0.033 | 9.2 | $1.29 \times 10^{-4}$ | intronic | *KIF16B* |

[1]Build hg38.

**Supplementary Table 6. *HIVEP3* variants associated with the SuperAger phenotype: comparison of summary statistics between SA vs. AD and SA vs. CU 80+ analyses**

| | | | | SuperAger vs. AD | | SuperAger vs. CU 80+ | |
|---|---|---|---|---|---|---|---|
| **Chr** | **ID** | **Position[1]** | **MAF** | **OR** | **P** | **OR** | **P** |
| 1 | rs12734651 | 41864914 | 0.30 | 0.24 | $6.46 \times 10^{-6}$ | 0.61 | 0.032 |
| 1 | rs6680922 | 41865541 | 0.30 | 0.24 | $7.35 \times 10^{-6}$ | 0.61 | 0.032 |
| 1 | rs7538225 | 41866890 | 0.30 | 0.24 | $7.37 \times 10^{-6}$ | 0.61 | 0.032 |
| 1 | rs6670896 | 41869122 | 0.30 | 0.24 | $7.32 \times 10^{-6}$ | 0.61 | 0.032 |
| 1 | rs6658462 | 41869277 | 0.30 | 0.24 | $7.32 \times 10^{-6}$ | 0.61 | 0.032 |
| 1 | rs12410469 | 41873116 | 0.30 | 0.24 | $7.32 \times 10^{-6}$ | 0.61 | 0.032 |
| 1 | rs11210536 | 41876744 | 0.30 | 0.24 | $7.71 \times 10^{-6}$ | 0.61 | 0.033 |
| 1 | rs11210537 | 41880052 | 0.30 | 0.24 | $7.64 \times 10^{-6}$ | 0.61 | 0.033 |

[1]Build hg38.

**Supplementary Table 7. *HIVEP3* variants associated with the SuperAger phenotype: comparison of summary statistics between SA vs. AD and SA vs. CU 80+ analyses (adjusted for *APOE* ε4 carrier status)**

| | | | | SuperAger vs. AD | | SuperAger vs. CU 80+ | |
|---|---|---|---|---|---|---|---|
| Chr | ID | Position[1] | MAF | OR | P | OR | P |
| 1 | rs12734651 | 41864914 | 0.30 | 0.26 | $1.99 \times 10^{-4}$ | 0.61 | 0.037 |
| 1 | rs6680922 | 41865541 | 0.30 | 0.26 | $2.03 \times 10^{-4}$ | 0.61 | 0.037 |
| 1 | rs7538225 | 41866890 | 0.30 | 0.26 | $2.03 \times 10^{-4}$ | 0.61 | 0.037 |
| 1 | rs6670896 | 41869122 | 0.30 | 0.26 | $2.02 \times 10^{-4}$ | 0.61 | 0.037 |
| 1 | rs6658462 | 41869277 | 0.30 | 0.26 | $2.02 \times 10^{-4}$ | 0.61 | 0.037 |
| 1 | rs12410469 | 41873116 | 0.30 | 0.26 | $2.02 \times 10^{-4}$ | 0.61 | 0.037 |
| 1 | rs11210536 | 41876744 | 0.30 | 0.26 | $2.21 \times 10^{-4}$ | 0.61 | 0.038 |
| 1 | rs11210537 | 41880052 | 0.30 | 0.26 | $2.19 \times 10^{-4}$ | 0.61 | 0.038 |

[1]Build hg38.

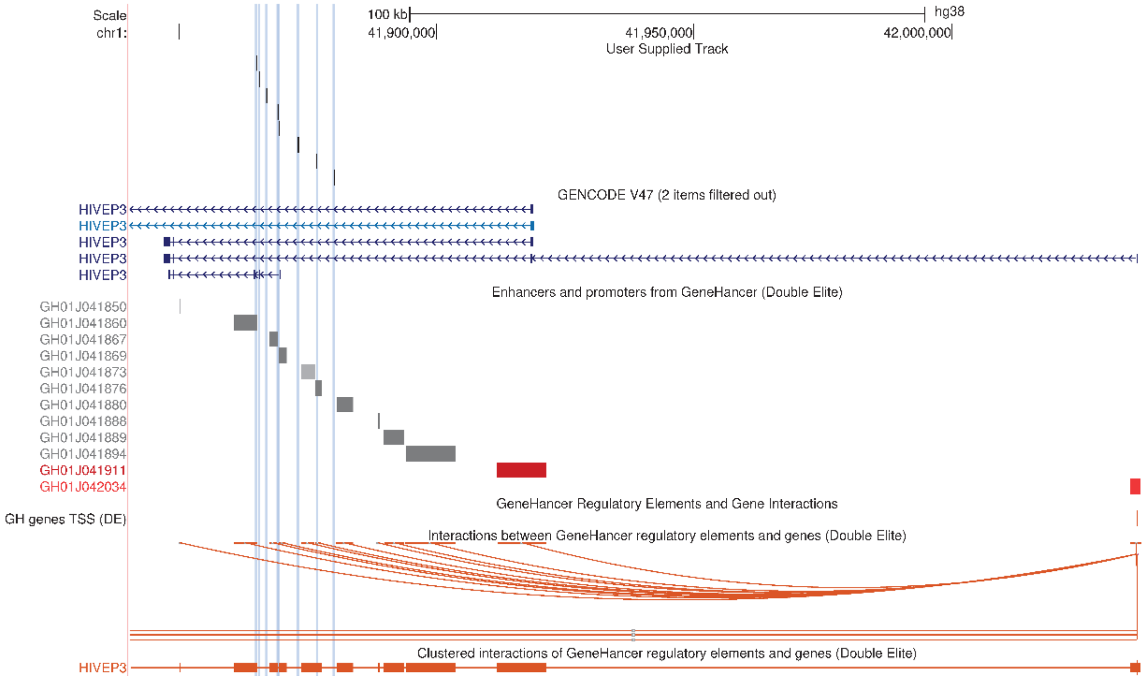


**Supplementary Figure 3. UCSC Genome Browser plot of *HIVEP3* locus**

The eight vertical blue lines correspond to the SNPs identified during the regional association screen. Beneath the transcript track are putative enhancer (grey) and promoter (red) elements as reported by GeneHancer.
